# The impact of hypertension and use of calcium channel blockers on tuberculosis treatment outcomes

**DOI:** 10.1101/2020.09.01.20185975

**Authors:** Vignesh Chidambaram, Akshay Gupte, Jann-Yuan Wang, Jonathan E. Golub, Petros C. Karakousis

**Affiliations:** Department of International Health, Johns Hopkins Bloomberg School of Public Health, Baltimore, Maryland; Center for Tuberculosis Research, Johns Hopkins University School of Medicine, Baltimore, MD, USA; Department of Internal Medicine, National Taiwan University Hospital, Taipei; Department of Epidemiology, Johns Hopkins Bloomberg School of Public Health; Department of Medicine, Johns Hopkins University School of Medicine, Baltimore, Maryland

**Keywords:** Tuberculosis, hypertension, calcium channel blockers, mortality, treatment outcomes

## Abstract

**Background:** Hypertension induces systemic inflammation, but its impact on the outcome of infectious diseases like tuberculosis (TB) is unknown. Calcium channel blockers (CCB) improve TB treatment outcomes in pre-clinical models, but their effect in patients with TB remain unclear.

**Methods:** This retrospective cohort study, including all patients > 18 years receiving treatment for culture-confirmed, drug-sensitive TB from 2000 to 2016 at the National Taiwan University Hospital, assessed the association of hypertension and CCB use with all-cause and infection-related mortality during the first 9 months of TB treatment, as well as sputum-smear microscopy and sputum-culture positivity at 2 and 6 months.

**Results:** 1052 of the 2894 patients (36.4%) had hypertension. Multivariable analysis revealed that hypertension was associated with increased mortality due to all causes (HR 1.57, 95% confidence interval[CI], 1.23-1.99) and infections (HR 1.87, 95%CI, 1.34-2.6), but there was no statistical difference in microbiological outcomes when stratified based on hypertensive group. Dihydropyridine-CCB (DHP-CCB) use was associated with reduced all-cause mortality (HR 0.67, 95%CI: 0.45-0.98) only by univariate Cox regression. There was no association between DHP-CCB use and infection-related mortality (HR 0.78, 95%CI: 0.46-1.34) or microbiological outcomes in univariate or multivariate regression analyses.

**Conclusions:** Patients with hypertension have increased all-cause mortality and infection-related mortality during the 9 months following TB treatment initiation. DHP-CCB use may lower all-cause mortality in TB patients with hypertension. The presence of hypertension or the use of CCB did not result in a significant change in microbiological outcomes.

## Introduction

Tuberculosis (TB) causes significant mortality worldwide, with approximately 1.5 million TB-related deaths in 2018, and nearly 90% of cases reported in developing countries [1]. Concurrently, the prevalence of hypertension has increased dramatically in many low- and middle-income countries, accounting for nearly 75% of global hypertension cases [2]. There is considerable overlap in countries with high burdens of TB and hypertension [3].

Patients with TB are significantly more likely to have hypertension than controls without TB[4]. Multiple cohort studies have shown an increased risk of cardiovascular and cerebrovascular disease following TB diagnosis [4–6]. Hypertension is associated with systemic inflammation, including increased levels of interleukin (IL)-1, IL-18, and tumor necrosis factor(TNF)-α. Although increased levels of pro-inflammatory mediators are associated with tissue destruction in pre-clinical models [7–9] and higher risk of advanced disease in patients with TB [10,11], the precise effect of hypertension on TB treatment outcomes is unknown.

Calcium channel blockers (CCB) are important first-line drugs used in the treatment of hypertension. Blockade of L-type voltage-gated calcium channels has been shown to improve mycobacterial clearance in preclinical studies [12]. Non-dihydropyridine CCB (non-DHP-CCB), such as verapamil, function as efflux pump inhibitors, decreasing the intrabacillary load in macrophages and increasing the bactericidal activity of bedaquiline against *Mycobacterium tuberculosis* in mouse models [13–16]. In contrast, dihydropyridine CCB (DHP-CCB), such as amlodipine or nifedipine, appear to lack efflux pump inhibitor activity. The effect of CCB use on TB treatment outcomes in humans is unknown.

In the current study, we assessed the impact of hypertension and CCB use on TB treatment outcomes in a retrospective cohort in Taiwan. Specifically, we evaluated the effect of hypertension and CCB use on 9-month all-cause mortality and infection-related mortality, as well as on sputum-culture positivity during standard treatment for drug-susceptible TB.

## Methods

### Study design and Population

We included all adults (aged >18 years) treated for drug-susceptible TB at the National Taiwan University Hospital (NTUH), a tertiary-care hospital situated in Taipei city, from 2000 to 2016. TB diagnosis was established by culture-based detection of *M. tuberculosis* in the sputum using MGIT-960 and Lowenstein–Jensen medium, and repeat sputum cultures were performed at 2 and 6 months after initiating TB treatment [17]. Most patients in this cohort were treated on an outpatient basis according to ATS guidelines [18]. There were no specific exclusion criteria. The study was approved by the institutional review boards at Johns Hopkins University and NTUH. All the data were obtained from the NTUH database.

### Baseline Characteristics

Data on age, sex, body mass index (BMI), diabetes mellitus (DM), cancer, chronic kidney disease (CKD), use of hemodialysis, cardiovascular disease, chronic obstructive pulmonary disease (COPD), asthma, liver disease, autoimmune disease, smoking, alcohol abuse, transplantation (both solid organ and bone marrow transplantation), HIV status, baseline sputum smear for acid-fast bacilli (AFB)(0 to 4+), cavitary disease on chest radiography, and prior history of TB were obtained.

### Exposures

The two exposures assessed in our study were 1) hypertension and 2) use of CCB among patients with hypertension. We defined hypertension at TB treatment initiation as systolic blood pressure > 140 mmHg or diastolic blood pressure >90 mmHg, according to the Joint National Committee-7 (JNC) and American Heart Association guidelines (2013) [19,20]. A minimum of 2 weeks (14 doses) of the following doses of CCB were taken as the required exposure: DHP-CCB [amlodipine (5mg), felodipine (5mg), lercanidipine (10mg), nifedipine (30mg), nicardipine (60mg)] or non-DHP-CCB [verapamil (240mg) and diltiazem (120 mg)]. CCB exposure was assigned separately for “intention-to-treat” and “per-protocol” analysis. Exposure for the “intention-to-treat” analysis was defined as any CCB use for at least 2 weeks in the first month of TB treatment and for the “per-protocol” analysis as the use of any CCB for > 80% (7.2 months) of the 9-month period following the initiation of TB treatment, or > 80% of the duration of follow-up if lost to follow-up or death occurred before 9 months. In order to appropriately assign CCB exposure, only the patients who survived beyond one month after TB treatment initiation were included for this analysis.

### Outcomes

The primary outcomes evaluated were all-cause and infection-related mortality during the first 9 months of TB treatment. The latter was a composite outcome of death due to pneumonia, sepsis, or TB during the first 9 months of TB treatment. Secondary outcomes included sputum-smear positivity and culture-confirmed TB at 2 and 6 months after treatment initiation.

### Statistical analysis

Participant characteristics stratified by hypertension and CCB exposure were compared using two-sided t-test and Chi-square test for continuous and categorical variables, respectively. Kaplan-Meier analysis and Cox regression was used to measure the association between the hypertension and all-cause and infection-related mortality in separate models. Person-time at risk of outcome stratified by hypertension status, was calculated from the time of TB treatment initiation until 9 months, or loss to follow-up or death, whichever occurred first. Sputum-smear and culture positivity at the end of 2 months were analysed using univariate and multivariate logistic regression. Similarly, using separate univariate and multivariate Cox regression models, we measured the association of CCB use, classified according to the “intention-to-treat” and “per-protocol” definitions, and the type of CCB (DHP-CCB or non-DHP-CCB), with all-cause mortality and infection-related mortality. In order to appropriately assign CCB exposure, only patients who survived beyond the first month post-TB treatment initiation were included. Sensitivity analyses were conducted by: i) including patients who died within the first month of TB treatment; and ii) excluding participants who did not receive the required dose of CCB from the comparison group. Potential confounders for multivariate analyses were identified by literature review and exploratory data analysis. Statistical analyses were performed using STATA/IC 16.0 software (StataCorp, College Station, Texas).

## RESULTS

### Stratification based on hypertension status

#### Sociodemographic characteristics and comorbidities of the study participants

Of the total 2894 culture-confirmed, drug-sensitive pulmonary TB cases, 1052 patients (36.4%) had hypertension (Table 1, Figure 1). The median (IQR) age was 66.6 years (49.1-77.8), with a higher proportion of patients aged>65 years in the hypertensive group (78.0%) relative to the normotensive group (37.7%) (p<0.001). The hypertensive group had higher percentage of males (72.2% vs 66.0%; p=0.862) and higher median BMI (22.0 vs 20.6; p=0.862) respectively, when compared to the normotensive group. Approximately 40% of the total patient population were ever-smokers, and 2.8% reported alcoholism, with no statistically significant difference between patients with and without hypertension.

**Table 1:**
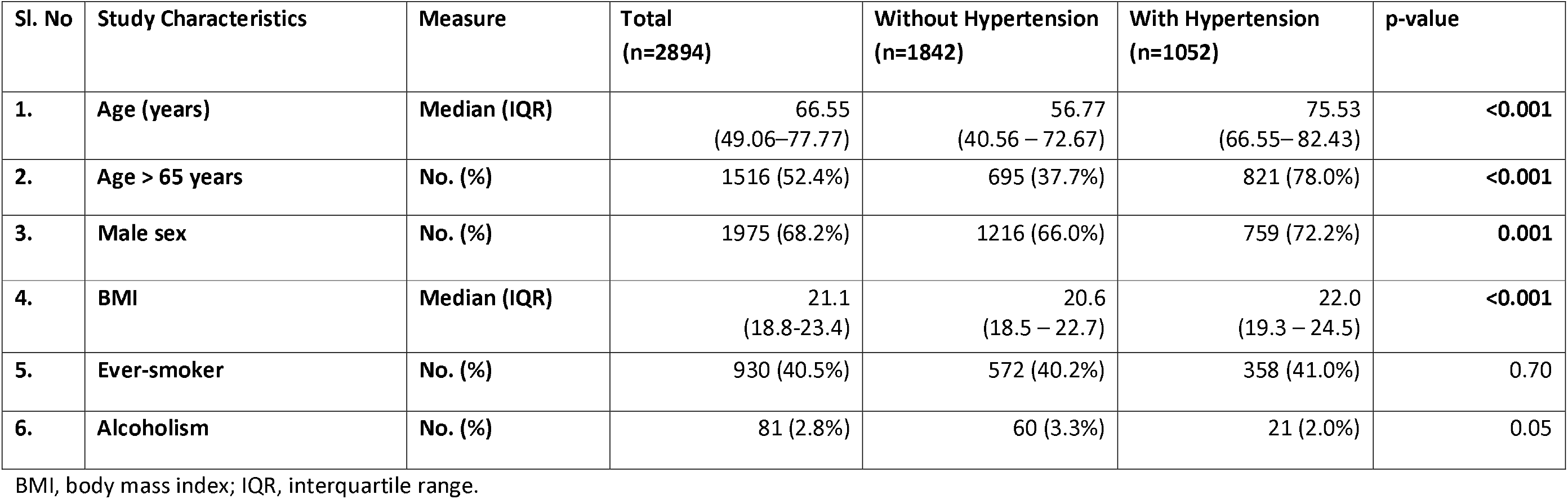
Sociodemographic characteristics of the study participants stratified by hypertension status (N=2894)

| Sl. No | Study Characteristics | Measure | Total<br>(n=2894) | Without Hypertension<br>(n=1842) | With Hypertension<br>(n=1052) | p-value |
| --- | --- | --- | --- | --- | --- | --- |
| 1. | Age (years) | Median (IQR) | 66.55<br>(49.06–77.77) | 56.77<br>(40.56 – 72.67) | 75.53<br>(66.55– 82.43) | <0.001 |
| 2. | Age > 65 years | No. (%) | 1516 (52.4%) | 695 (37.7%) | 821 (78.0%) | <0.001 |
| 3. | Male sex | No. (%) | 1975 (68.2%) | 1216 (66.0%) | 759 (72.2%) | 0.001 |
| 4. | BMI | Median (IQR) | 21.1<br>(18.8-23.4) | 20.6<br>(18.5 – 22.7) | 22.0<br>(19.3 – 24.5) | <0.001 |
| 5. | Ever-smoker | No. (%) | 930 (40.5%) | 572 (40.2%) | 358 (41.0%) | 0.70 |
| 6. | Alcoholism | No. (%) | 81 (2.8%) | 60 (3.3%) | 21 (2.0%) | 0.05 |

BMI, body mass index; IQR, interquartile range.

**Figure 1:**
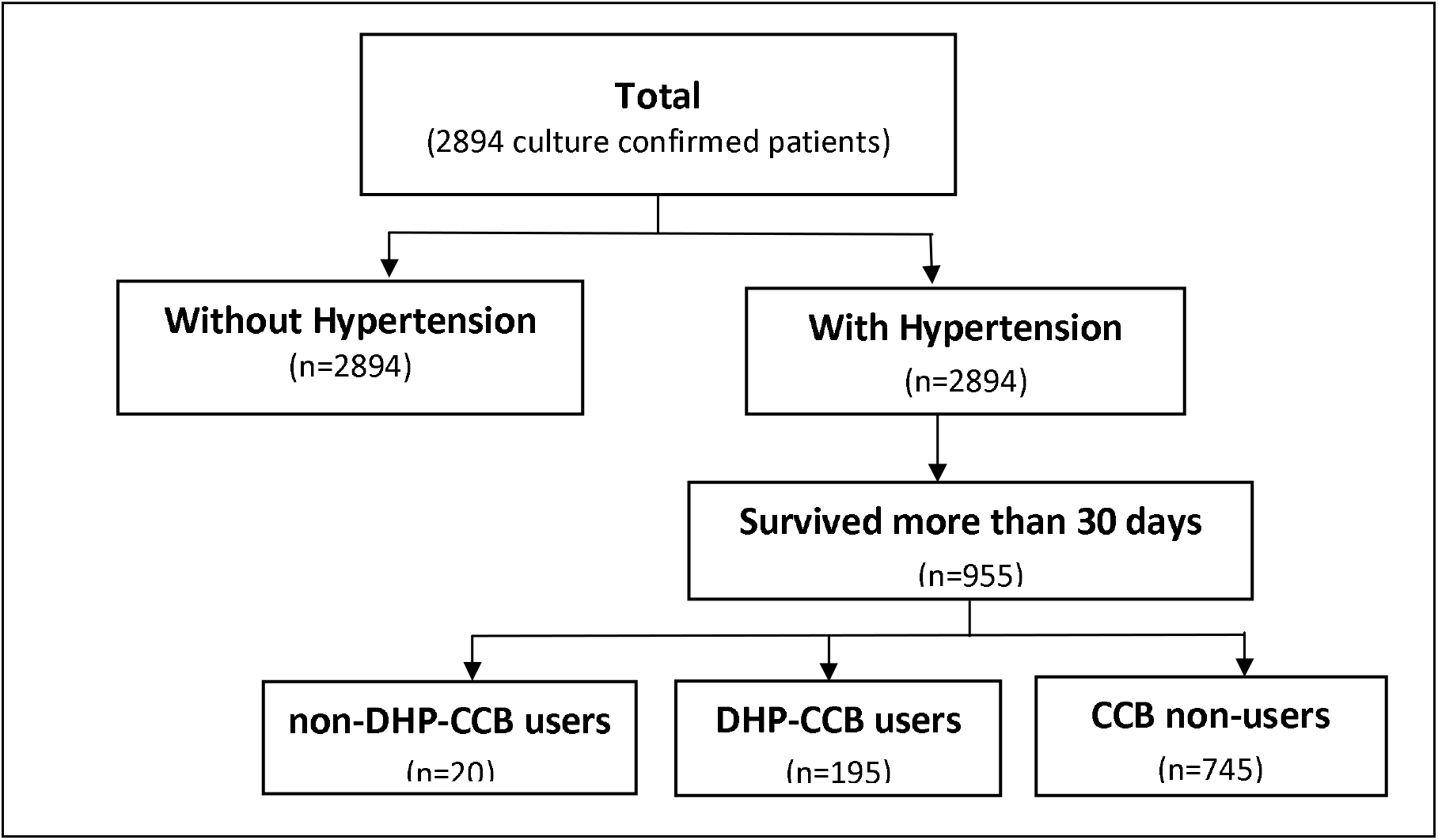
Flow chart for participant selection

There were higher rates of DM, cardiovascular disease, peripheral vascular disease, cerebrovascular accidents, cancer, CKD, COPD, and cirrhosis among patients in the hypertensive group (Table 2). A greater proportion of patients being smear-positive by microscopy at diagnosis in the normotensive group (45.8%) compared to the hypertensive group (37.4%) (p<0.001) (Table 3). A higher percentage of patients in the normotensive group (16.6%) had cavitary disease compared to the hypertensive group (10.3%) (p<0.001).

**Table 2:** Associated co-morbidities of the study participants stratified by hypertension status (N=2894)

| Sl. No | Study Characteristics | Measure | Total<br>(n=2894) | Without Hypertension<br>(n=1842) | With Hypertension<br>(n=1052) | p-value |
| --- | --- | --- | --- | --- | --- | --- |
| 1. | DM | No. (%) | 533 (18.5%) | 212 (11.5%) | 321(30.6%) | <0.001 |
| 2. | Cancer | No. (%) | 459(15.9%) | 238(12.9%) | 221(21.1%) | <0.001 |
| 3. | CKD stage ≥ 4 | No. (%) | 128/2513(5.1%) | 24/1519(1.6%) | 104/994(10.5%) | <0.001 |
| 4. | Cardiovascular disease | No. (%) | 352 (12.1%) | 98 (5.3%) | 254 (24.1%) | <0.001 |
|  | CAD | No. (%) | 291 (10.0%) | 81 (4.4%) | 210 (19.9%) | <0.001 |
|  | AMI | No. (%) | 111 (3.8%) | 46 (2.5%) | 65 (6.2%) | <0.001 |
|  | CHF | No. (%) | 123 (4.2%) | 28 (1.5%) | 95 (9.00%) | <0.001 |
|  | PVD | No. (%) | 83 (2.9%) | 28 (1.5%) | 55 (5.2%) | <0.001 |
| 5. | Asthma | No. (%) | 123 (4.2%) | 69 (3.7%) | 54 (5.1%) | 0.07 |
| 6. | COPD | No. (%) | 451 (15.6%) | 223 (12.1%) | 228 (21.7%) | <0.001 |
| 7. | Bronchiectasis | No. (%) | 121 (4.2%) | 82 (4.5%) | 39 (3.7%) | 0.34 |
| 8. | Pneumoconiosis | No. (%) | 35 (1.2%) | 22 (1.2%) | 13 (1.2%) | 0.92 |
| 9. | Autoimmune diseases | No. (%) | 115 (4.0%) | 64 (3.5%) | 51 (4.9%) | 0.07 |
| 10. | Cirrhosis | No. (%) | 50 (1.7%) | 29 (1.6%) | 21 (2.0%) | 0.40 |
| 11. | History of transplant | No. (%) | 26 (0.9%) | 10 (0.5%) | 16 (1.5%) | 0.007 |
| 12. | HIV | No. (%) | 65 (2.3%) | 54 (2.9%) | 11 (1.1%) | 0.001 |
| 13. | Charlson Comorbidity index | Mean (SD) | 3.9 (2.7) | 3.0 (2.4) | 5.4 (2.4) | <0.001 |

Abbreviations: AMI, Acute myocardial infarction; CAD, Coronary artery disease; CHF, Congestive Heart failure; CKD, Chronic kidney disease; COPD, Chronic obstructive pulmonary disease; DM, diabetes mellitus; HIV, Human Immunodeficiency Virus; PVD, Peripheral Vascular Disease. \*Denominators are mentioned when they are different from the "n"

**Table 3:** TB disease characteristics of the study participants (N=2894)

| Sl. No | Study Characteristics | Measure | Total (n=2894) | Without Hypertension (n=1842) | With Hypertension (n=1052) | p-value |
| --- | --- | --- | --- | --- | --- | --- |
| 1. | Sputum Smear AFS positive at baseline | No. (%) | 1215/2842 (42.8%) | 831/1817 (45.8%) | 384/1025 (37.4%) | < 0.001 |
| 2. | Sputum Smear AFS grade at baseline |  |  |  |  | <0.001 |
|  | 0 |  | 1627/2842 (57.3%) | 986/1817 (54.3%) | 641/1029 (39.4%) |  |
|  | 1+ |  | 390/2842 (13.7%) | 246/1817 (13.5%) | 144/1029 (14.1%) |  |
|  | 2+ |  | 318/2842 (11.2%) | 228/1817 (12.6%) | 90/1029 (8.8%) |  |
|  | 3+ |  | 241/2842 (8.5%) | 164/1817 (9.0%) | 77/1029 (7.5%) |  |
|  | 4+ |  | 266/2842 (9.4%) | 193/1817 (10.6%) | 73/1029 (7.1%) |  |
| 3. | Prior TB | No. (%) | 98/1598 (6.1%) | 65/1040 (6.3%) | 33/558 (5.9%) | 0.79 |
| 4. | Cavity | No. (%) | 413 (14.3%) | 305 (16.6%) | 108 (10.3%) | <0.001 |

AFS, Acid fast smear. TB, Tuberculosis. \*Denominators are mentioned when they are different from the “n”

### Outcomes

#### Effect of hypertension on 9-month all-cause mortality

During the first 9 months after TB treatment initiation, there was a significantly higher all-cause mortality in the hypertensive group (290/997 patients, 29.1%) than in the non-hypertensive group (254/1670 patients, 15.2%) (p<0.001) (Table 4). The log-rank test of the Kaplan-Meier analysis showed that patients with hypertension had significantly shorter survival compared to patients without hypertension (Figure 2A; p< 0.001). After adjusting for confounders, patients with hypertension had a HR of 1.57(95%CI 1.23-1.99) for all-cause mortality during TB treatment (Table 5). Other parameters found to be associated with increased 9-month all-cause mortality in the adjusted Cox-regression models were male sex, lower BMI, sputum AFB smear positivity and higher CCI score.

**Table 4:** Outcomes in patients with TB stratified by hypertension status (N=2894)

| Sl. No | Study Characteristics | Measure | Total<br>(n=2894) | Without Hypertension<br>(n=1842) | With Hypertension<br>(n=1052) | p-value |
| --- | --- | --- | --- | --- | --- | --- |
| 1. | All-cause mortality within 9 months | No. (%) | 544/2667<br>(20.4%) | 254/1670<br>(15.2%) | 290/997<br>(29.1%) | <0.001 |
| 2. | Infection-related mortality within 9 months | No. (%) | 303/2667<br>(11.4%) | 135/1670<br>(8.1%) | 168/997<br>(16.9%) | <0.001 |
| 3. | Sputum culture positive at 2 months | No. (%) | 265/1640<br>(16.2%) | 165/1020<br>(16.2%) | 100/620<br>(16.1%) | 0.980 |
| 4. | Sputum AFB smear positive at 2 months | No. (%) | 118/1640<br>(7.2%) | 81/1020<br>(7.9%) | 37/620<br>(6.0%) | 0.13 |
| 5. | Sputum culture positive at 6 months | No. (%) | 6/1551<br>(0.4%) | 5/978<br>(0.5%) | 1/573<br>(0.2%) | 0.30 |
| 6. | Sputum AFB smear positive at 6 months | No. (%) | 1/1551<br>(0.1%) | 1/978<br>(0.1%) | 0/573<br>(0%) | 0.44 |

AFB, Acid fast bacilli. \*Denominators are mentioned when they are different from the “n”

**Figure 2A:**
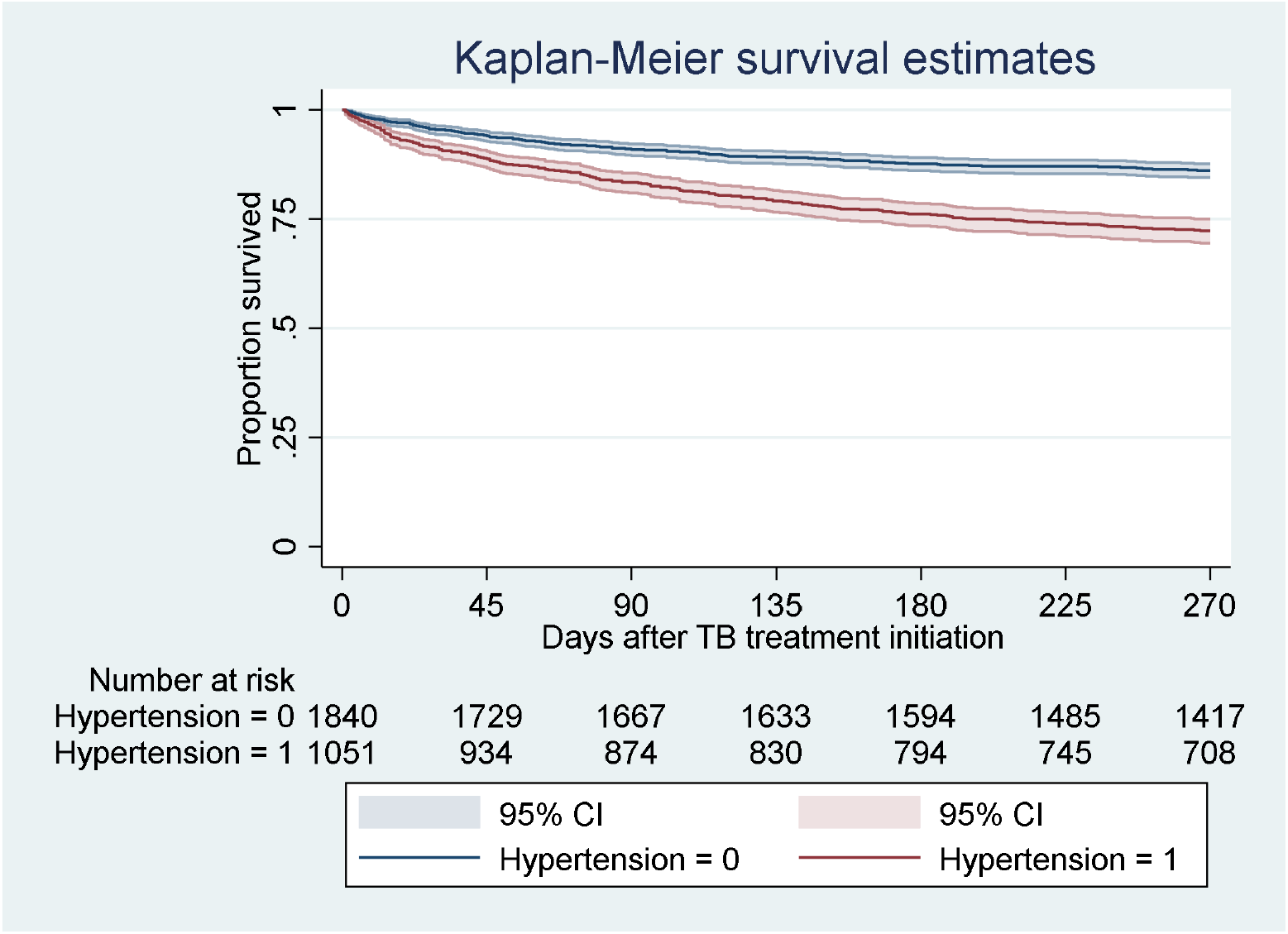
9-month survival after TB treatment initiation among patients with (1) and without (0) hypertension.

**Table 5:**
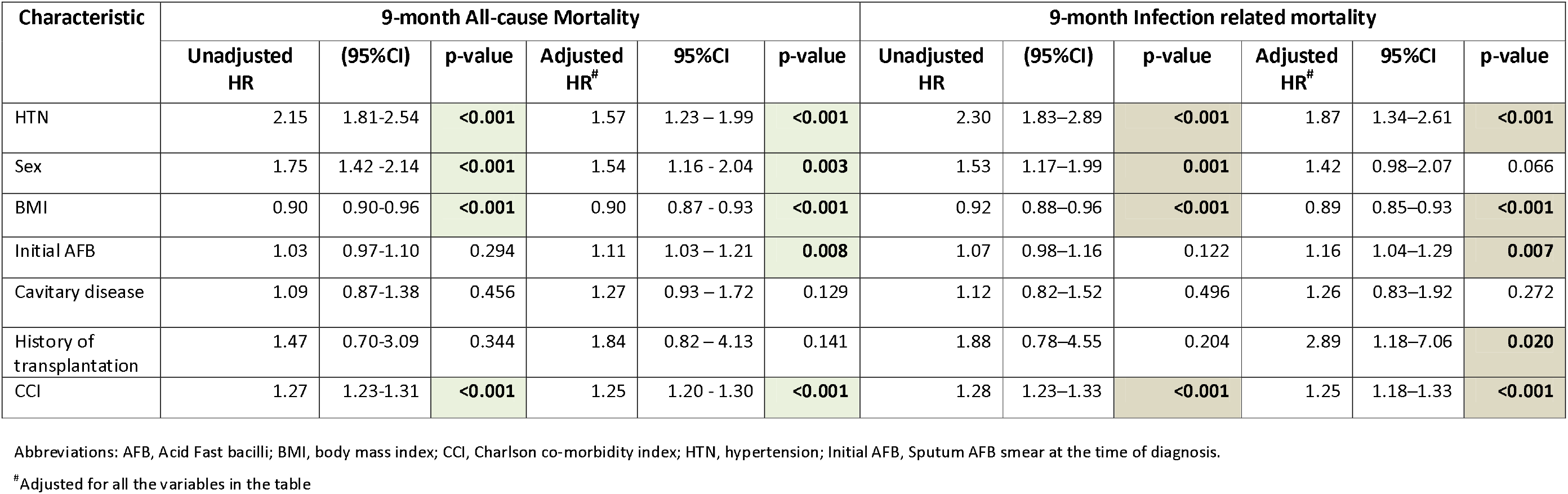
Crude and adjusted hazard ratios, based on Cox regression model, for all-cause and infection-related mortality during 9 months after initiation of TB treatment (N = 2915)

#### Effect of hypertension on 9-month infection-related mortality

During the first 9 months after TB treatment initiation, a total of 303/2667 patients (11.4%) died of infection-related causes, constituting 55.7% (303/544 patients) of all deaths during the first 9 months of TB treatment (Table 4). Of this group, 43 patients (14.2%) died due to TB-related causes, 124 patients (40.9%) due to pneumonia and 136 patients (44.9%) due to sepsis. Hypertension was associated with significantly earlier infection-related mortality by the log-rank test of the Kaplan-Meier analysis (Figure 2B; p< 0.001). After adjusting for confounders, patients with hypertension had 1.87 times higher hazard of mortality due to infection during TB treatment (95% CI, 1.34–2.61; P < 0.001).

**Figure 2B:**
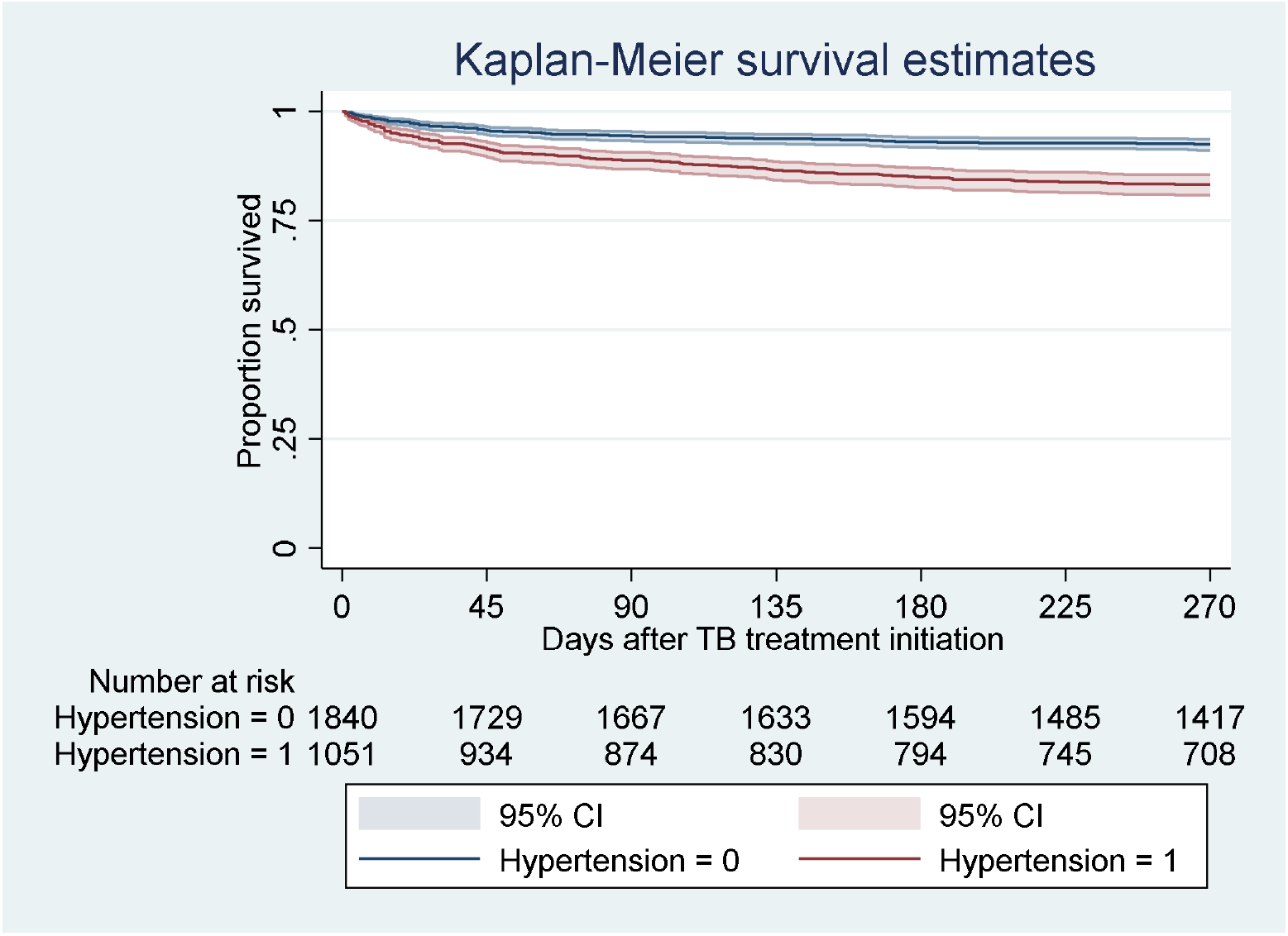
Survival analysis for 9-month infection-related mortality after TB treatment initiation among patients with (1) and without (0) hypertension.

#### Effect of hypertension on microbiological outcomes

Among the 2915 patients included in our study, 265/1640 patients (16.2%) and 6/1551 patients (0.4%) had positive sputum cultures at 2 months and 6 months, respectively. There were no differences among the hypertensive and normotensive groups with respect to the proportion of sputum-culture positivity either at 2 months (16.1% vs 16.2%, respectively; p=0.862) or 6 months (0.2% vs 0.6%, respectively; p=0.216) (Table 4). Univariate and multivariate models of logistic regression also showed no significant relationship between hypertension and sputum culture or microscopy positivity at 2 months (Supplementary Table 1A).

### Stratification based on CCB use

Among the 955 hypertensive patients with follow-up more than 30 days after TB diagnosis, 195 patients (20.4%) received DHP-CCB and 20 patients (2.1%) received non-DHP-CCB, while 5 patients (0.5%) received both DHP-CCB and non-DHP-CCB. Of the 955 patients with hypertension, 54 (5.6%) of these patients were lost to follow up during the first 9 months of treatment. The results of the per-protocol analyses are described in Table 7. Baseline characteristics among CCB users and non-users were not statistically different, except that the CCB use group included a higher percentage of patients with prior TB (13.3% vs 5.2%) (p<0.01).

#### Stratification of hypertensive patients based on CCB use

Of the total 955 patients with hypertension, 210 patients (77.9%) received the required dose of either CCB type (DHP or non-DHP). Among them, 32/201 patients (16.0%) died of any cause and 29/187 patients (15.4%) died of infection-related causes. Univariate Cox-regression shows a hazard ratio (HR) of 0.68 (95%CI: 0.47-0.99) for all-cause mortality in patients who received any CCB (Table 8). Multivariate analysis after controlling for prior TB history did not yield a significant association with all-cause mortality. Intention-to-treat analysis did not reveal a significant difference in time to all-cause mortality between CCB users and non-users (Supplementary Table 2). Univariate and multivariate analysis did not yield a significant association for infection-related mortality during the first 9 months of TB treatment. Chi-square test (Table 7) and univariate and multivariate logistic regression (Supplementary table 1B) revealed no association between CCB use and microbiological outcomes.

#### Stratification of hypertensive patients based on non-DHP-CCB use

Twenty patients (1.6%) satisfied the criteria for non-DHP-CCB use. There was no statistically significant difference in all-cause mortality between non-DHP-CCB use (4/15 patients) and CCB non-use (162/704 patients) (26.67% vs 20.79%) (p=0.852). Similarly, no statistical difference was found in infection-related mortality between the non-DHP-CCB group (3/15 patients) and non-CCB group (72/704 patients) (20.0% vs 10.80%) (p=0.314).

#### Stratification of hypertensive patients based on DHP-CCB use

As shown in Table 6, there were more male patients (p=0.03) and patients with a prior history of TB (p<0.01) in patients receiving DHP-CCB compared to those not receiving CCB. Patients receiving DHP-CCB had significantly lower mortality (29/187, 15.2%) compared to those not treated with CCB (162/701, 23.1%; p=0.02, Table 7). Kaplan-Meier survival analysis and log-rank test shows that patients treated with DHP-CCB had a significantly longer survival compared to patients not receiving DHP-CCB (Figure 3A; p-value= 0.041), while multivariate Cox regression did not show a statistically significant relationship to all-cause mortality (Table 8). Intention-to-treat analysis did not reveal a significant difference in time to all-cause mortality between DHP-CCB users and CCB non-users (Supplementary Table 2). There were no statistically significant differences in 9-month infection-related mortality (Figure 3B), or sputum-culture conversion rates at 2 and 6 months of TB treatment among patients with and without DHP-CCB use when chi-square test (Table 7), and univariate and multivariate logistic regression were used (Supplementary Table 1B).

**Table 6:** Baseline characteristics of participants with hypertension stratified by CCB use (per-protocol) (N=959)

| Sl. No | Study Characteristics | Measure | Total (n=959) | All CCB use |  |  | DHP-CCB use |  |  |
| --- | --- | --- | --- | --- | --- | --- | --- | --- | --- |
|  |  |  |  | Without CCB use (n=745) | With CCB use (n=210) | p-value | Without CCB use (n=745) | With DHP-CCB use (n=195) | p-value |
| 1. | Age (years) | Median (IQR) | 75.1 (66.4 –82.0) | 75.1 (66.2-82.2) | 75.0 (67.0-80.8) | 0.36 | 75.1 (66.2-82.2) | 74.7 (66.7– 80.6) | 0.57 |
| 2. | Male Sex | No. (%) | 71.94% | 73.4% | 66.7% | 0.05 | 73.4% | 65.6% | 0.03 |
| 3. | Initial AFB smear | No. (%) | 348 (36.7%) | 271 (36.6%) | 77 (36.8%) | 0.95 | 271 (36.6%) | 72 (37.1%) | 0.90 |
| 5. | Prior TB | No. (%) | 32 (6.8%) | 19 (5.2%) | 13 (13.3%) | <0.01 | 19 (5.2%) | 12 (13.0%) | <0.01 |
| 6. | Cavitory disease | No. (%) | 98 (10.3%) | 75 (10.1%) | 23 (11.0%) | 0.71 | 75 (10.1%) | 22 (11.3%) | 0.62 |
| 7. | DM | No. (%) | 287 (30.1%) | 220 (29.6%) | 67 (31.9%) | 0.52 | 220 (29.6%) | 62 (31.8%) | 0.55 |
| 8. | Cancer | No. (%) | 194 (20.4%) | 153 (20.6%) | 41 (19.5%) | 0.73 | 153 (20.6%) | 40 (20.5%) | 0.98 |
| 9. | CKD Stage ≥ 4 | No. (%) | 85 (9.4%) | 65 (9.1%) | 20 (10.5%) | 0.58 | 65(9.1%) | 19 (10.8%) | 0.51 |
| 10. | Cardiovascular disease | No. (%) | 213(22.2%) | 166 (22.2%) | 47 (22.3%) | 0.98 | 166(22.2%) | 41 (20.9%) | 0.63 |
|  | CAD |  | 181 (18.9%) | 142 (19.0%) | 39 (18.5%) | 0.87 | 142(19.0%) | 34 (17.4%) | 0.61 |
|  | AMI |  | 54 (5.6%) | 44 (5.9%) | 10 (4.7%) | 0.53 | 44(5.9%) | 9 (4.6%) | 0.48 |
|  | CHF |  | 71 (7.4%) | 59 (7.9%) | 12 (5.7%) | 0.28 | 59(7.9%) | 10 (5.1%) | 0.18 |
| 11. | Asthma | No. (%) | 49 (5.1%) | 37 (5.0%) | 12 (5.7%) | 0.67 | 37(5.0%) | 9 (4.6%) | 0.84 |
| 12. | COPD | No. (%) | 214 (22.4%) | 157 (21.1%) | 57 (27.1%) | 0.06 | 157 (21.1%) | 53 (27.2%) | 0.07 |
| 13. | Bronchiectasis | No. (%) | 39 (4.1%) | 26 (3.5%) | 13 (6.2%) | 0.08 | 26(3.5%) | 12 (6.1%) | 0.09 |
| 14. | Pneumoconiosis | No. (%) | 13 (1.4%) | 10 (1.3%) | 3 (1.4%) | 0.93 | 10(1.3%) | 2 (1.0%) | 0.73 |
| 15. | Autoimmune disease | No. (%) | 44 (4.6%) | 31 (4.1%) | 13 (6.2%) | 0.22 | 31(4.1%) | 13 (6.7%) | 0.14 |
| 16. | Liver cirrhosis | No. (%) | 17 (1.8%) | 14 (1.8%) | 3 (1.5%) | 0.66 | 14(1.8%) | 3 (1.5%) | 0.75 |
| 17. | History of transplant | No. (%) | 16 (1.7%) | 12 (1.6%) | 4 (1.9%) | 0.77 | 12 (1.6%) | 4 (2.0%) | 0.67 |
| 18. | HIV | No. (%) | 9 (0.9%) | 7 (0.9%) | 2 (1.0%) | 0.99 | 7(0.9%) | 2 (1.0%) | 0.91 |
| 19. | CCI | Mean (SD) | 5.2 (2.3) | 5.2 (2.3) | 5.4 (2.1) | 0.25 | 5.2 (2.3) | 5.4 (2.1) | 0.16 |

Abbreviations: AFB, Acid Fast bacilli; AMI, Acute myocardial infarction; BMI, body mass index; CAD, Coronary artery disease; CHF, Congestive Heart failure; CKD, Chronic kidney disease; COPD, Chronic obstructive pulmonary disease; DM, diabetes mellitus; HIV, Human Immunodeficiency Virus; MDR-Tb, Multi drug resistant TB
A Ninety-seven patients were excluded from this analysis as they had a follow up time of less than 30 days since TB diagnosis.

**Table 7:** Outcomes in patients with hypertension stratified by DHP-CCB use (N=2915)

| Sl. No | Study Characteristics | Measure | Total (n=959) | All CCB use |  |  | DHP-CCB use |  |  |
| --- | --- | --- | --- | --- | --- | --- | --- | --- | --- |
|  |  |  |  | Without CCB use (n=745) | With CCB use (n=210) | p-value | Without CCB use (n=745) | With DHP-CCB use (n=195) | p-value |
| 1. | All-cause mortality within 9 months | No. (%) | 194/905 (21.4%) | 162/701 (23.1%) | 32/201 (16.0%) | 0.03 | 162/701 (23.1%) | 29/187 (15.2%) | 0.03 |
| 2. | Infection-related mortality within 9 months | No. (%) | 106/905 (11.7%) | 86/704 (12.3%) | 20/201 (10.0%) | 0.38 | 86/704 (12.2%) | 18/188 (9.6%) | 0.11 |
| 3. | Sputum AFB smear positive at 2 months | No. (%) | 36/616 (5.8%) | 32/474 (6.8%) | 4/142 (2.8%) | 0.08 | 32/474 (6.6%) | 4/131 (3.1%) | 0.11 |
| 4. | Sputum AFB smear positive at 6 months | No. (%) | 0/569 (0%) | 0/443 (0%) | 0/126 (0%) | - | 0/443 (0%) | 0/117 (0%) | - |
| 5. | Sputum culture positive at 2 months | No. (%) | 99/616 (16.1%) | 75/474 (15.8%) | 24/142 (16.8%) | 0.76 | 77/474 (15.8%) | 22/131 (16.8%) | 0.79 |
| 6. | Sputum culture positive at 6 months | No. (%) | 1/569 (0.2%) | 0/443 (0%) | 1/126 (0.2%) | 0.59 | 0/443 (0%) | 1/117 (0.2%) | 0.607 |

Abbreviations: AFB, Acid Fast bacilli; CCB, Calcium channel blocker; DHP-CCB, Di-hydropyridine calcium channel blocker.
\*Denominators are mentioned when they are different from the "n"

**Figure 3A:**
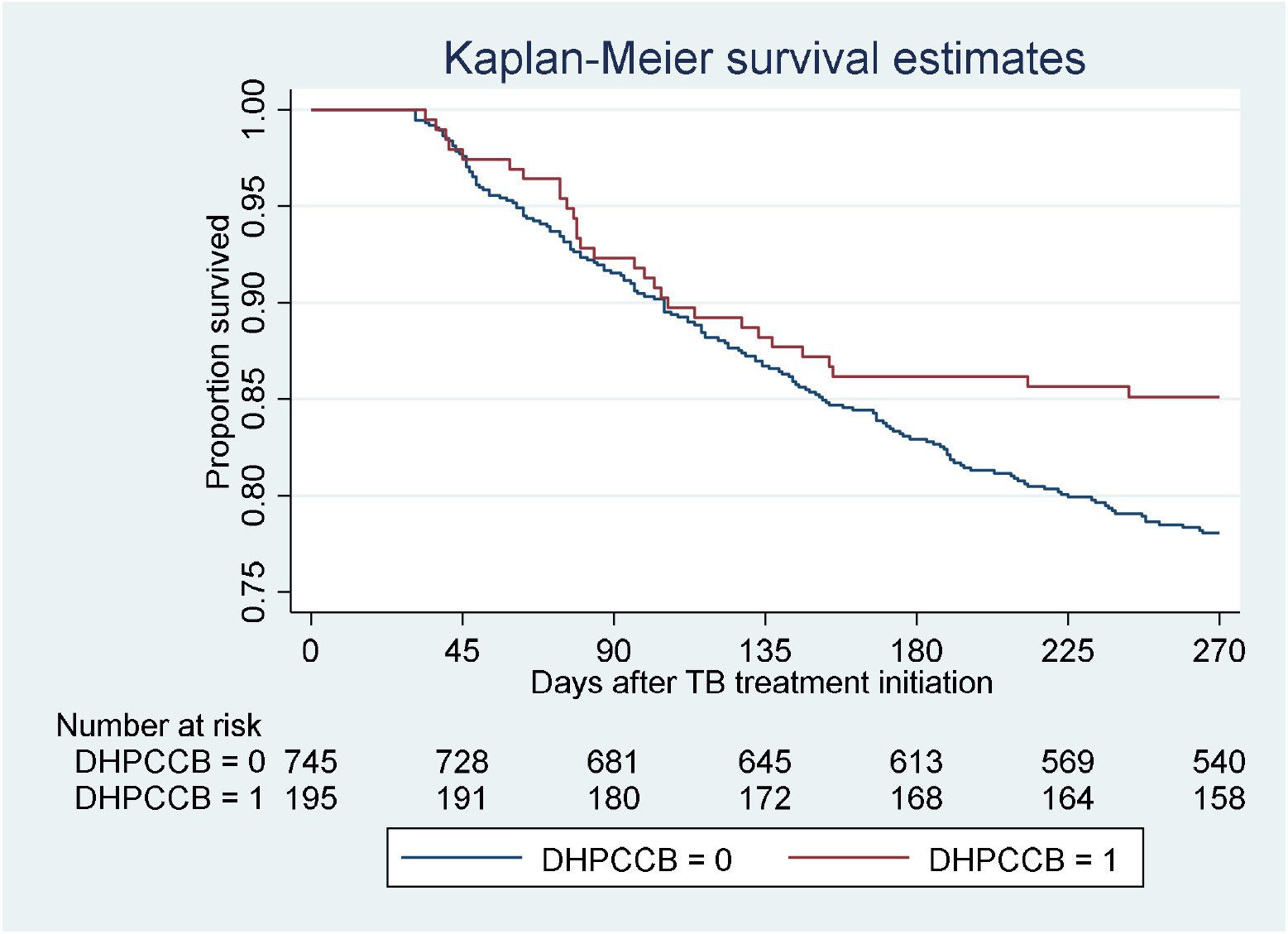
9-month all-cause mortality following TB treatment initiation among DHP-CCB users

**Table 8:**
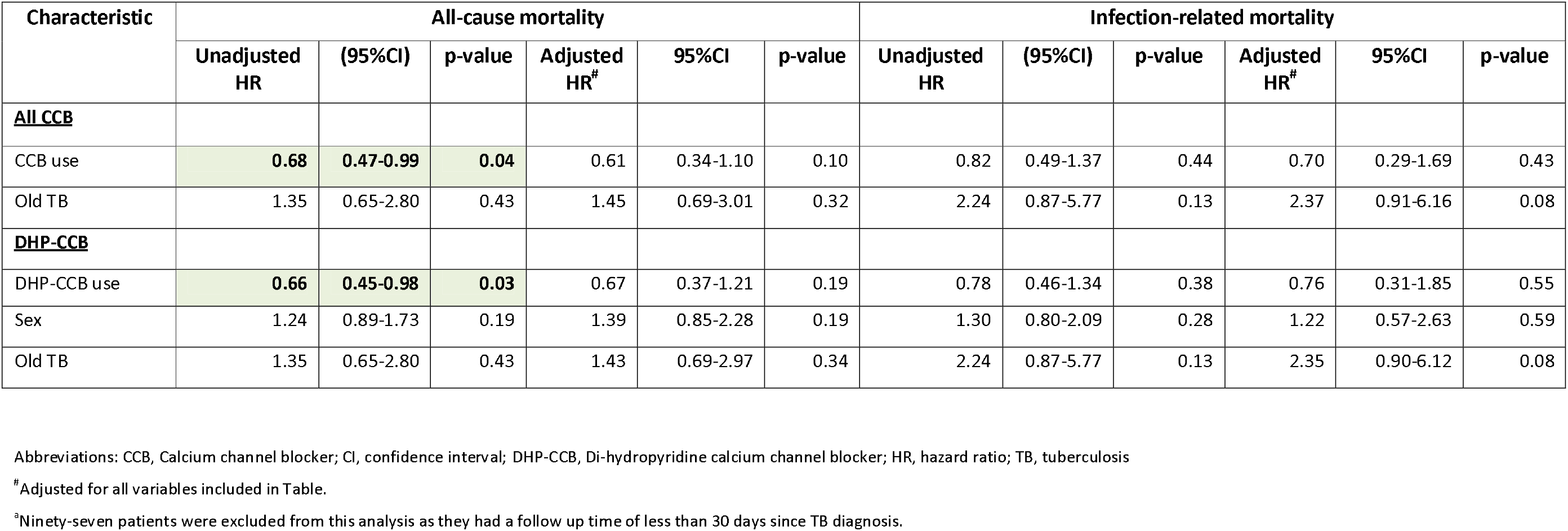
Crude and adjusted hazard ratios, based on Cox regression model, for all-cause mortality and infection-related mortality during 9 months after initiation of TB treatment among patients with hypertension and CCB use (per-protocol analysis) (N = 2915)

| Characteristic | All-cause mortality |  |  |  |  |  | Infection-related mortality |  |  |  |  |  |
| --- | --- | --- | --- | --- | --- | --- | --- | --- | --- | --- | --- | --- |
|  | Unadjusted HR | (95%CI) | p-value | Adjusted HR<sup>#</sup> | 95%CI | p-value | Unadjusted HR | (95%CI) | p-value | Adjusted HR<sup>#</sup> | 95%CI | p-value |
| **<u>All CCB</u>** |  |  |  |  |  |  |  |  |  |  |  |  |
| CCB use | **0.68** | **0.47-0.99** | **0.04** | 0.61 | 0.34-1.10 | 0.10 | 0.82 | 0.49-1.37 | 0.44 | 0.70 | 0.29-1.69 | 0.43 |
| Old TB | 1.35 | 0.65-2.80 | 0.43 | 1.45 | 0.69-3.01 | 0.32 | 2.24 | 0.87-5.77 | 0.13 | 2.37 | 0.91-6.16 | 0.08 |
| **<u>DHP-CCB</u>** |  |  |  |  |  |  |  |  |  |  |  |  |
| DHP-CCB use | **0.66** | **0.45-0.98** | **0.03** | 0.67 | 0.37-1.21 | 0.19 | 0.78 | 0.46-1.34 | 0.38 | 0.76 | 0.31-1.85 | 0.55 |
| Sex | 1.24 | 0.89-1.73 | 0.19 | 1.39 | 0.85-2.28 | 0.19 | 1.30 | 0.80-2.09 | 0.28 | 1.22 | 0.57-2.63 | 0.59 |
| Old TB | 1.35 | 0.65-2.80 | 0.43 | 1.43 | 0.69-2.97 | 0.34 | 2.24 | 0.87-5.77 | 0.13 | 2.35 | 0.90-6.12 | 0.08 |

Abbreviations: CCB, Calcium channel blocker; CI, confidence interval; DHP-CCB, Di-hydropyridine calcium channel blocker; HR, hazard ratio; TB, tuberculosis
<sup>#</sup>Adjusted for all variables included in Table.
<sup>a</sup>Ninety-seven patients were excluded from this analysis as they had a follow up time of less than 30 days since TB diagnosis.

**Figure 3B:**
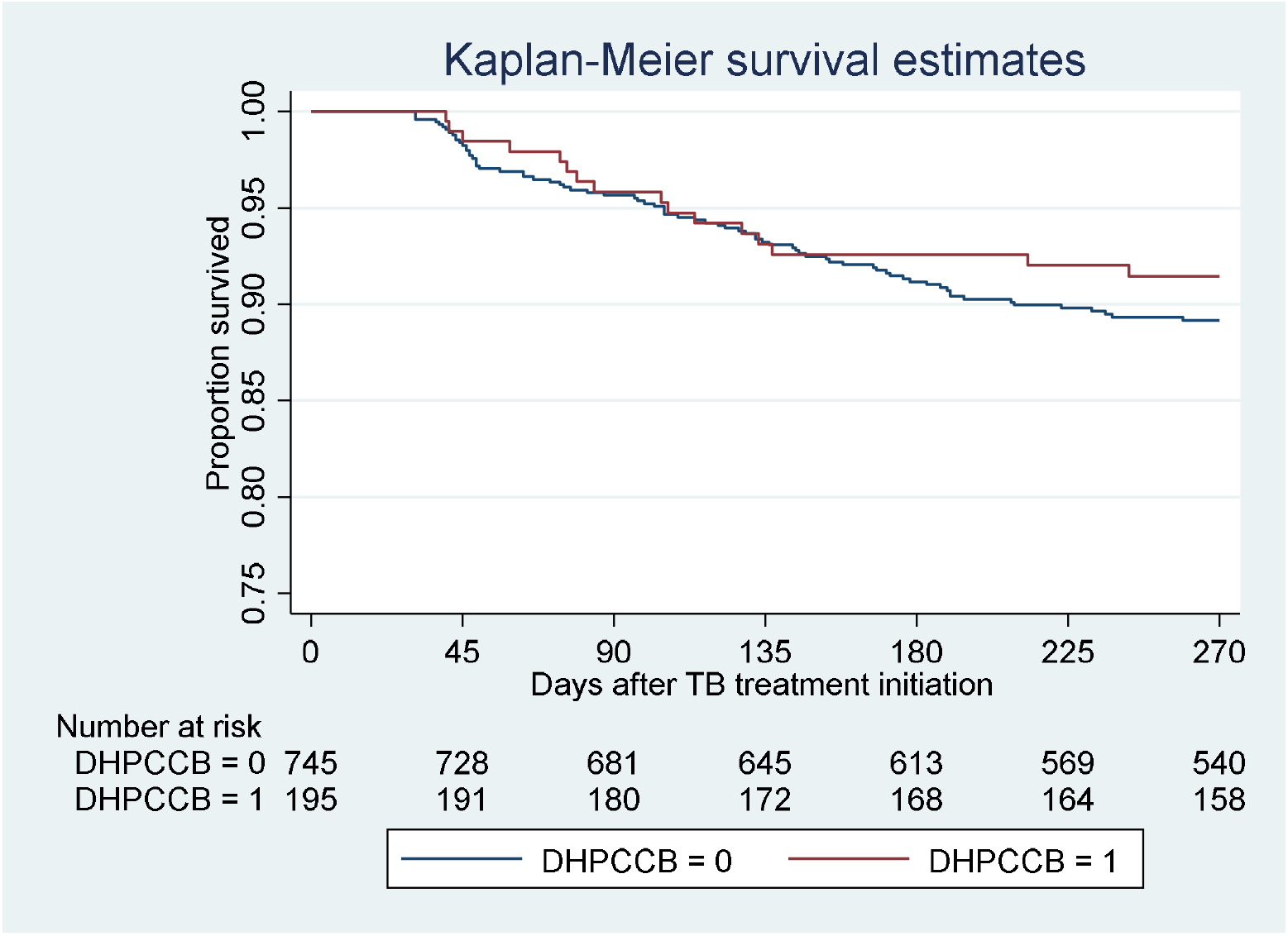
9-month infection-related mortality following TB treatment initiation among DHP-CCB users

#### Sensitivity analysis

Sensitivity analysis performed by including patients who died in the first month of TB treatment also showed a lower hazard of all-cause mortality with any CCB use (HR 0.68, 95%CI 0.47-0.99) and DHP-CCB use (HR 0.48, 95%CI 0.33-0.70). However, multivariate Cox regression revealed no significant association with all-cause mortality, and infection-related mortality in patients with hypertension was not found to be associated with CCB use (Supplementary Table 3A). Sensitivity analysis by excluding participants who did not receive a required dose of CCB did not show any significant association with all-cause mortality or infection-related mortality (Supplementary Table 3B).

## DISCUSSION

In our cohort of TB patients, 821/1516 patients (54.2%) aged more than 65 years had hypertension. This proportion is considerably higher than that reported previously by Umeki *et al*. in a similar population in Japan (35%) [21]. Of note, most patients with hypertension at the time of TB diagnosis do not report a history of hypertension [22]. Therefore, our findings would suggest that screening for hypertension is important, especially in elderly patients receiving a new TB diagnosis.

The major finding of our study is that patients with hypertension had a greater adjusted hazard of all-cause mortality (HR 1.57, 95%CI 1·23–1.99) and, interestingly, infection-related mortality (HR 1.87, 95%CI 1·34–2·61) during the first 9 months following initiation of TB treatment. In a prior study by Chiang *et al*., although hypertension was associated with an increase in the composite outcome of treatment failure, default, and death [23], it was not shown to be an independent predictor of negative outcomes when adjusted for other factors [24]. In that study, death constituted a very small proportion (17.3%) of negative outcomes and a much greater proportion was contributed by treatment default (78.2%) [24]. Our study is the first to show increased short-term mortality attributable to hypertension in patients receiving TB treatment. We found that hypertension was an independent predictor of 9-month all-cause and infection-related mortality, when adjusting for sex, BMI, sputum acid-fast stain at the time of diagnosis, cavitary disease, past history of transplantation, and CCI scores. As age is an important component of CCI, this finding raises the possibility that there could be a direct link connecting hypertension and poor TB treatment outcomes, apart from the age-related co-morbidities frequently observed in patients with hypertension.

Hypertension has long been recognized as an inflammatory condition [25]. Increased levels of C-reactive protein, TNF-α and T-helper cells were observed in patients with hypertension compared to non-hypertensive patients [26,27]. Animal models have shown that decreased levels of renin and angiotensin were associated with decreased levels of peripheral T-lymphocytes and interleukins, such as interleukin IL-1α, IL-1β, and IL-6 [28]. Hypertension is also associated with increased oxidative stress [29] and angiotensin II is known to increase the production of reactive oxygen species through nicotinamide adenosine dinucleotide phosphate (NADPH) oxidase [30]. The relationship between NADPH oxidase and ROS with sepsis-induced mortality has been shown in a mouse model by Kong *et al*. [31]. The higher risk of infection-related mortality seen in hypertensive patients in our study could be due to the increased levels of interleukins, NADPH oxidase, and reactive oxygen species in these patients [32].

While a number of other infections, such as Aggregatibacter, Prevotella [33], *Helicobacter pylori* [34], HIV [35–37], herpes simplex virus-2 [38], cytomegalovirus [39], *Toxoplasmosis gondii* [40] have been shown to be associated with an increased risk of hypertension, there are no prior studies which assessed the outcome of these infections in the hypertensive patients. In our study, the proportion of patients who had sputum AFB smear positivity and cavitary disease at the time of diagnosis was higher in the non-hypertensive group compared to the hypertensive group, suggesting a higher bacillary burden in the lungs of patients in the former group. However, at 2 and 6 months after TB treatment initiation, there was no statistically significant difference by Chi-square tests and multivariable logistic regression, in rates of sputum AFB smear positivity between the two groups. This suggests that hypertension could interfere with mycobacterial clearance. Other factors associated with worse clinical outcomes at 9 months of TB treatment were the presence of cardiovascular disease, CKD stage ≥ 4 and the presence of cancer. These are consistent with the available evidence from the other retrospective cohort studies performed in patients with TB [17,42].

Blockade of the L-type and R-type voltage-gated calcium channels in macrophages, which are responsible for regulating phagocytic function, have been shown to increase intracellular calcium concentration [12]. This, in turn, has been demonstrated to increase the killing of intracellular mycobacteria in mouse models [12]. Verapamil, a non-DHP-CCB which blocks mycobacterial efflux pumps, has been shown to reduce TB treatment time and decrease the minimum inhibitory concentration of bedaquiline and clofazimine in pre-clinical studies [43]. In addition, verapamil inhibits host cell efflux pumps, and has attracted attention as a potential host-directed therapy [14,15]. In our study, only 20 patients with hypertension satisfied the criteria for exposure to verapamil and other non-DHP-CCB. Among these patients, there was no significant difference in the 9-month all-cause mortality during TB treatment (p=0.852). A clinical trial with a larger sample size of patients receiving non-DHP-CCB is required to further evaluate the potential adjunctive activity of this class of agents in TB treatment. There are no previous studies performed using DHP-CCB as an adjunctive therapy. Our study revealed that DHP-CCB use in hypertensive patients surviving more than 30 days following the initiation of TB treatment was associated with a significant decrease in 9-month all-cause mortality. This finding could not be explained by differences in baseline characteristics between patients receiving and those not receiving DHP-CCB. Interestingly, there was a non-statistically significant trend towards lower sputum AFB positivity rate at 2 months in patients who used DHP-CCB (3.1%) compared to those who did not (6.6%). The culture conversion rates at 2 and 6 months were not statistically significant between the DHP-CCB users and the DHP-CCB non-users.

An important limitation of our study is that sputum smear microscopy and microbiological culture results at 2 months and 6 months post-TB treatment were not available for nearly 25% of the patients in our cohort. There was a drop-out rate of 9%, which could be a source of bias in our study. There were inconsistencies in the reporting of cause of death, which prevented us from accurately assessing TB-specific mortality. The level of control of hypertension and other anti-hypertensive drugs were not obtainable from the available data, which could be confounding factors in study. The availability of precise systolic and diastolic pressure data could permit the evaluation of a possible dose-response curve between these parameters and mortality, as well as CCB blocker efficacy and study outcomes. On the other hand, the large sample size of our cohort enabled the analysis of individual risk factors and their impact on short-term mortality. Additionally, enrolment of all the study participants from a single centre helped to decrease heterogeneity in the clinical management of these patients.

Our study shows that hypertension is an independent predictor of all-cause mortality during the first 9 months after the initiation of TB treatment after adjustment for sex, BMI, initial sputum AFB smear, history of transplantation and cavitary disease. Also, hypertension was associated with a higher incidence of 9-month infection-related mortality, which included TB-related deaths, pneumonia, and sepsis. Microbiological outcomes were not significantly different between the hypertensive and non-hypertensive groups, despite a higher incidence of smear-positive and cavitary disease in the non-hypertensive group at baseline. The use of DHP-CCB was associated with improved 9-month all-cause mortality in hypertensive patients with TB. Our study highlights the importance of screening for hypertension, especially in elderly patients receiving a new diagnosis of TB. Prospective studies are warranted to determine the potential beneficial effects of controlling hypertension, and specifically use of CCB, on TB treatment outcomes.

## Data Availability

Data will be made available at request to the corresponding author

